# Patient outcomes after hospitalisation with COVID-19 and implications for follow-up; results from a prospective UK cohort

**DOI:** 10.1101/2020.08.12.20173526

**Authors:** DT Arnold, FW Hamilton, A Milne, A Morley, J Viner, M Attwood, A Noel, S Gunning, J Hatrick, S Hamilton, KT Elvers, C Hyams, A Bibby, E Moran, H Adamali, J Dodd, NA Maskell, S Barratt

**Affiliations:** Academic Respiratory Unit, North Bristol NHS Trust, Southmead Way, Bristol, BS105NB; Bristol Centre for Antimicrobial Research (BCARE), North Bristol NHS Trust, Southmead Way, Bristol, BS105NB; Medicines Discovery Institute Cardiff, Cardiff University, Park Place, Cardiff, CF10 3AT

**Author notes:** Joint first authorship (DT Arnold, FW Hamilton). Corresponding author: Dr David Arnold, Academic Respiratory Unit, Learning and Research Centre, Southmead Hospital, Southmead Way, BS10 5NB.

## Abstract

**Background:** COVID-19 causes a wide spectrum of disease. The incidence and severity of sequelae after the acute infection is uncertain. Data measuring the longer-term impact of COVID-19 on symptoms, radiology and pulmonary function are urgently needed to plan follow-up services.

**Methods:** Consecutive patients hospitalised with COVID-19 were prospectively recruited to this observational study with outcomes recorded at 28-days. All were invited to a systematic follow up at 8-12 weeks, including chest radiograph, spirometry, exercise test, bloods, and health-related quality of life (HRQoL) questionnaires.

**Findings:** Between 30^th^ March and 3^rd^ June 2020, 163 patients with COVID-19 were recruited. Median hospital length of stay was 5 days (IQR 2-8) and 19 patients died. At 8-12 weeks post admission, 134 patients were available for follow up and 110 attended. Most (74%) had persistent symptoms (notably breathlessness and excessive fatigue) with reduced HRQoL.

Only patients who required oxygen therapy in hospital had abnormal radiology, clinical examination or spirometry at follow up. Thirteen (12%) patients had an abnormal chest X-ray with improvement in all but 2 from admission. Eleven (10%) had restrictive spirometry. Blood test abnormalities had returned to baseline in the majority (104/110).

**Interpretation:** Patients with COVID-19 remain highly symptomatic at 8-12 weeks, however, clinical abnormalities requiring action are infrequent, especially in those without a supplementary oxygen requirement during their acute illness. This has significant implications for physicians assessing patients with persistent symptoms, suggesting that a more holistic approach focussing on rehabilitation and general wellbeing is paramount.

**Funding:** Southmead Hospital Charity

## BACKGROUND

The effect of SARS-CoV-2 varies dramatically between individuals from asymptomatic infection through to respiratory dysfunction and multi-organ failure. In the United Kingdom (UK) over 130,000 people have been hospitalised with COVID-19, with 17% requiring intensive care admission and a 26% mortality rate.^1 2^ Treatment strategies for the acute phase of COVID-19 have been investigated with several effective agents identified.^3 4^ However, very limited data exists on the medium-term outcomes of patients who survived. Such data will allow the design of appropriate follow-up protocols in a healthcare system with large waiting lists and limitations on face-to-face appointments.

Several guidelines have been published on the appropriate medium and longer term follow-up of patients who survived COVID-19.^5-7^ Given the lack of evidence these are based on extrapolation of longer-term data from other coronavirus infections.^8 9^ It is vital that disease specific data on the outcomes for survivors of COVID-19, both on symptomatology and clinical tests, is used to properly inform guidelines.

We report the first prospectively recruited UK cohort of hospitalised patients with COVID-19. Consecutively hospitalised patients were recruited at diagnosis and outcomes assessed at 28 days. Participants were then followed-up at 8-12 weeks with face-to-face medical review, spirometry, exercise test, bloods, chest radiograph and assessment of physical and mental health-related quality of life (HRQoL). We aimed to assess the prevalence of complications from COVID-19 within these patients to inform appropriate follow up in secondary or primary care.

## METHODS

### Subjects

All patients were recruited via the DISCOVER study (Diagnostic and Severity markers of COVID-19 to Enable Rapid triage study), a single centre prospective study recruiting consecutive patients admitted with COVID-19, from 30.03.2020 until present (Ethics approval via South Yorkshire REC: 20/YH/0121, CRN approval no: 45469). The inclusion criteria were a positive PCR result for SARS-CoV-2, using the established Public Health England reverse transcriptase polymerase chain reaction (RT-PCR) assay in use at the time, or a clinico-radiological diagnosis of COVID-19 disease. The only exclusion criteria were age <18 and the inability to give informed consent to study participation. All patients gave verbal consent (witnessed by 2 members of the study team) for storage of routinely collected clinical data and research blood samples. For patients in respiratory high care or intensive care who were too unwell to provide consent, family members were contacted to provide a declaration as a personal consultee, in line with the framework under the Mental Capacity Act (2005).

### Baseline assessment

Routine demographics were recorded including ethnicity, and presence of important comorbidities. The earliest admission National Early Warning Score (NEWS) was extracted from the clinical record. This is a numeric score (from 1-20), reflecting the degree of physiological dysfunction.^10^ Routine biochemistry and haematology results were extracted from the clinical record (C-reactive protein (CRP), neutrophils, lymphocytes, neutrophil:lymphocyte ratio), using the admission results. Chest radiography was performed on admission and radiological severity score calculated (see below).

### 28-day remote follow-up

All recruited patients were followed up remotely at 28-days after admission by review of hospitals notes and/or general practice records. This included 28-day mortality, hospital length of stay, readmissions, requirement for intensive care, ventilation, renal replacement therapy, and inotropes. We also recorded complications including acute renal failure, acute liver injury, venous thromboembolic events (both pulmonary emboli and deep vein thromboses), cardiac events (including myocardial infarction, myocarditis, congestive cardiac failure and arrythmias), and neurological events (cerebrovascular events, meningitis or encephalitis).

At 28 days, participants were defined as having had severe disease (death, requirement for non-invasive ventilation (NIV), intensive care or high dependency unit admission), moderate disease (requirement for oxygen during hospital stay), or mild disease (no requirement for oxygen or enhanced care during stay).

### 8-12 week face-to-face outpatient follow up

All patients who survived were invited to a follow up at a respiratory outpatient clinic (with the exception of nursing home residents or current hospital inpatients). All patients who attended this appointment had a face-to-face review with a respiratory or infectious disease clinician, chest radiograph, spirometry, exercise testing, routine bloods, routine observations and HRQoL questionnaires (see details below).

#### Chest radiograph

Non-portable radiography equipment was used to obtain posterior-anterior (PA) projection radiographs with standard techniques at a 180-cm focus-film distance.

The radiological severity score was calculated for the baseline radiograph using the method described by Wong et al, 2020.^11^ A score of 0-4 was assigned to each lung depending on the extent of involvement by consolidation or ground glass opacities. 0 = no involvement, 1 = <25%, 2 = 25 - 49%, 3= 50 - 75%, 4 = >75% involvement. The scores for each lung were summed to produce a final severity score ranging from 0-8. Radiographs were scored by a respiratory and infectious diseases physician.

All follow-up chest radiographs were categorised into two groups, normal or abnormal, based on lung parenchymal, airway, pleural, hilar and mediastinal findings as reported by a consultant radiologist and verified by the attending respiratory physician.

In those chest x-rays demonstrating an abnormality, the lung parenchyma and airways were evaluated for the following: 1) consolidation, 2) ground-glass opacity (GGO), 3) nodular opacity, and 4) reticular opacity 5) atelectasis 6) pleural pathology, by consultant radiologists and according to standardised terminology.^12^

#### Spirometry

Forced expiratory volume during first second of expiration (FEV1) and forced vital capacity (FVC) were performed in accordance with ATS/ERS guidelines.^13^ The MRC score, height (meters), and body weight of the patients (kilograms) were also recorded.^14^ Lung physiology staff wore full personal protective equipment (PPE) during testing including FFP-3 masks.

#### Sit to stand test (STS)

Given the importance of social distancing in clinical areas, exercise testing was assessed using the 1-min sit-to-stand test (STS) as opposed to the 6-minute walk test (6MWT). All 1-min STS tests were performed according to a standardised protocol using a standard chair (height 46-48 cm) with a flat seat and no armrests. Patients were instructed to stand completely straight from a seated position and touch the chair with their bottom when sitting, but that they need not sit fully back on the chair. Patients were asked to complete the manoeuvre without using their hands or arms to assist movement and to perform as many repetitions as possible in 1 min. A minimum of three sit to stands were required in order for this to be recoded as an adequate test. The resting oxygen saturation was recorded via pulse oximetry, in addition to the nadir oxygen saturation during the test and up to one minute during recovery. A mild desaturation was classified as ≥ 4% but with a nadir ≥94%, a significant desaturation was classified as a ≥ 4% desaturation but with a nadir <94%.^15^

#### Health status questionnaires

The SF-36 is a questionnaire of 36 items, measuring eight multi-item variables; physical functioning (PF), social functioning (SF), role limitations due to physical (RP) or emotional problems (RE), mental health (MH), energy and vitality (VT), bodily pain (BP) and general perception of health (GH).^16^ There is a further single item for perception of change in health over the past year. For each variable, items are scored and transformed into a scale of 0 to 100 (best possible health status). Subsequently, a composite physical and mental composite score (PCS, MCS) are generated from each individual variant. The WEMWBS is a scale of 14 positively worded items for assessing feeling and functioning aspects of mental wellbeing, with 5 response categories of ‘none of the time’ ‘to ‘all of the time’. Scores ranged from 14-70, with higher scores indicating greater positive mental wellbeing.^17^

### Statistical analysis

Categorical variables were presented as counts with percentages. All continuous data were non-parametric and therefore presented with medians and interquartile range (IQR), unless otherwise specified. Differences between patient groups were evaluated using Mann Whitney-U and Kruskal Wallis tests for continuous data and Fisher’s exact test or Chi-squared testing for categorical data. Statistical significance was taken as p ≤ 0.05. Data were analysed using R version 4.0.0 with the packages “tidyverse” and “gtsummary”.

## FINDINGS

Between 30^th^ March and 3^rd^ June 2020, 163 participants with COVID-19 were recruited to this study. The median age of the cohort was 60 (IQR: 46-73) and 91 (56%) were male. Nineteen patients died and 13 were either nursing home residents or current inpatients at the time of planned follow-up. We invited the remaining 131 for follow-up via telephone or letter 8-12 weeks after their hospital admission. Eighteen declined follow-up due to ongoing shielding (n=10), being care-providers (n=3) or feeling follow-up was unnecessary (n=5), and 3 were not-contactable. Therefore 110 patients attended hospital follow-up. There were no significant differences in medical history or clinical factors between patients who attended versus those who did not attend follow-up (see Supplementary material). For subsequent analyses, only the followed-up cohort is described. Figure 1 shows the CONSORT diagram of the study. Table 1 shows the baseline demographics and clinical outcomes of those who attended follow-up divided by severity of COVID-19 illness. Patients were followed up a median of 83 days (IQR 74-88 days) after hospital admission and 90 days (IQR 80-97 days) after onset of their COVID-19 symptoms.

**Figure 1:**
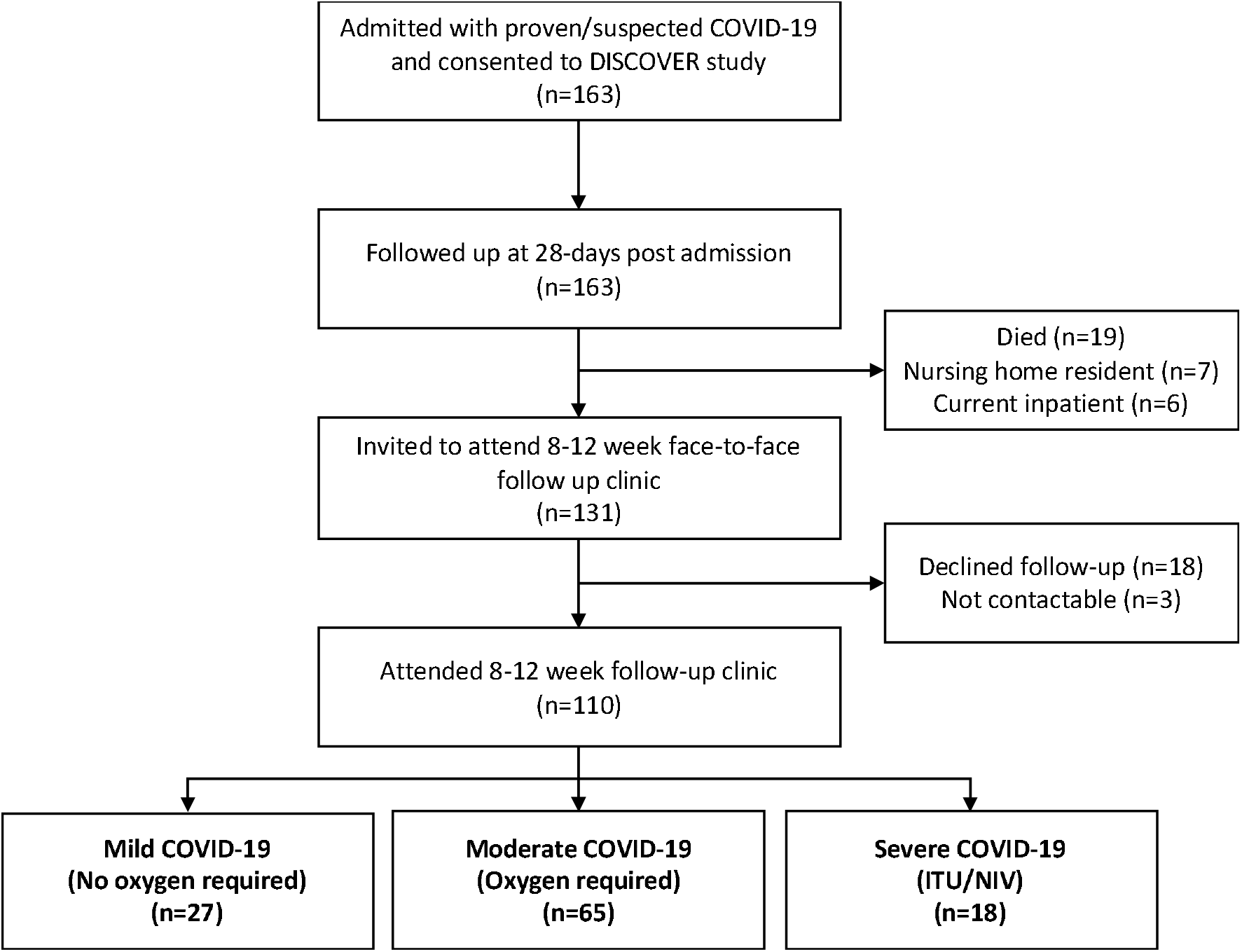
CONSORT diagram of the study.

**Table 1:** Demographics and admission factors of patients attending follow-up (n=110)

| Characteristic | Mild<br>(n = 27) | Moderate<br>(n = 65) | Severe<br>(n = 18) |
| --- | --- | --- | --- |
| *Demographics* |  |  |  |
| Age (18+) | 47 (32, 61) | 57 (48, 67) | 62 (54, 71) |
| BAME | 5 (19%) | 15 (23%) | 3 (19%) |
| Male | 13 (48%) | 44 (68%) | 11 (61%) |
| BMI (mean) | 31.2 | 32.5 | 32.5 |
| *Co-morbidities* |  |  |  |
| T1DM | 1 (3.7%) | 1 (1.5%) | 1 (5.6%) |
| T2DM | 2 (7.4%) | 12 (18%) | 2 (11%) |
| Heart disease | 6 (22%) | 11 (17%) | 3 (17%) |
| Chronic Lung disease | 4 (15%) | 16 (25%) | 8 (44%) |
| Severe Liver disease | 0 (0%) | 1 (1.5%) | 0 (0%) |
| Severe kidney disease | 1 (3.7%) | 4 (6.2%) | 2 (11%) |
| Hypertension | 4 (15%) | 16 (25%) | 7 (39%) |
| HIV | 0 (0%) | 0 (0%) | 1 (5.6%) |
| *Laboratory Testing* |  |  |  |
| SARS CoV-2 PCR +ve<br>(as inpatient) | 21 (78%) | 50 (77%) | 10 (56%) |
| SARS-CoV-2-IgG +ve (Abbott)<br>(at follow-up) | 18 (67%) | 56 (86%) | 15 (83%) |
| *Outcomes* |  |  |  |
| Admission (ED) NEWS score (IQR) | 2 (1, 3) | 4 (2, 6) | 5 (4, 8) |
| Radiographic severity score on<br>admission chest radiograph | 0 (0-2) | 3 (1.75-4) | 3 (1.25 -6) |
| Invasive or non-invasive ventilation<br>required | 0 (0%) | 0 (0%) | 16 (89%) |
| Supplementary oxygen required | 0 (0%) | 65 (100%) | 18 (100%) |
| Hospital length of stay (days) | 2 (1-4) | 5 (2-8) | 10 (7-17) |
| *BAME- Black, Asian and Minority Ethnic, BMI-Body Mass Index, DM- Diabetes mellitus, HIV- Human Immunodeficiency Virus, NEWS- National Early Warning Score.* |  |  |  |

### Symptoms

Symptoms were common at follow up, with 81 (74%) patients reporting at least one ongoing symptom since discharge from hospital. Sixteen (59%) patients in the mild COVID-19 group reported ongoing symptoms compared to 49 (75%) and 16 (89%) in the moderate and severe groups respectively. The frequency of symptoms reported at follow-up compared to the point of hospital admission is shown in Figure 2. The most common symptoms at follow-up were breathlessness, excessive fatigue (39% prevalence each) and insomnia (24%), with the incidence of insomnia apparently increased at follow-up compared to baseline. Patients with more severe disease were more symptomatic especially in terms of breathlessness, fatigue, myalgia and insomnia, see Supplementary material.

**Figure 2:**
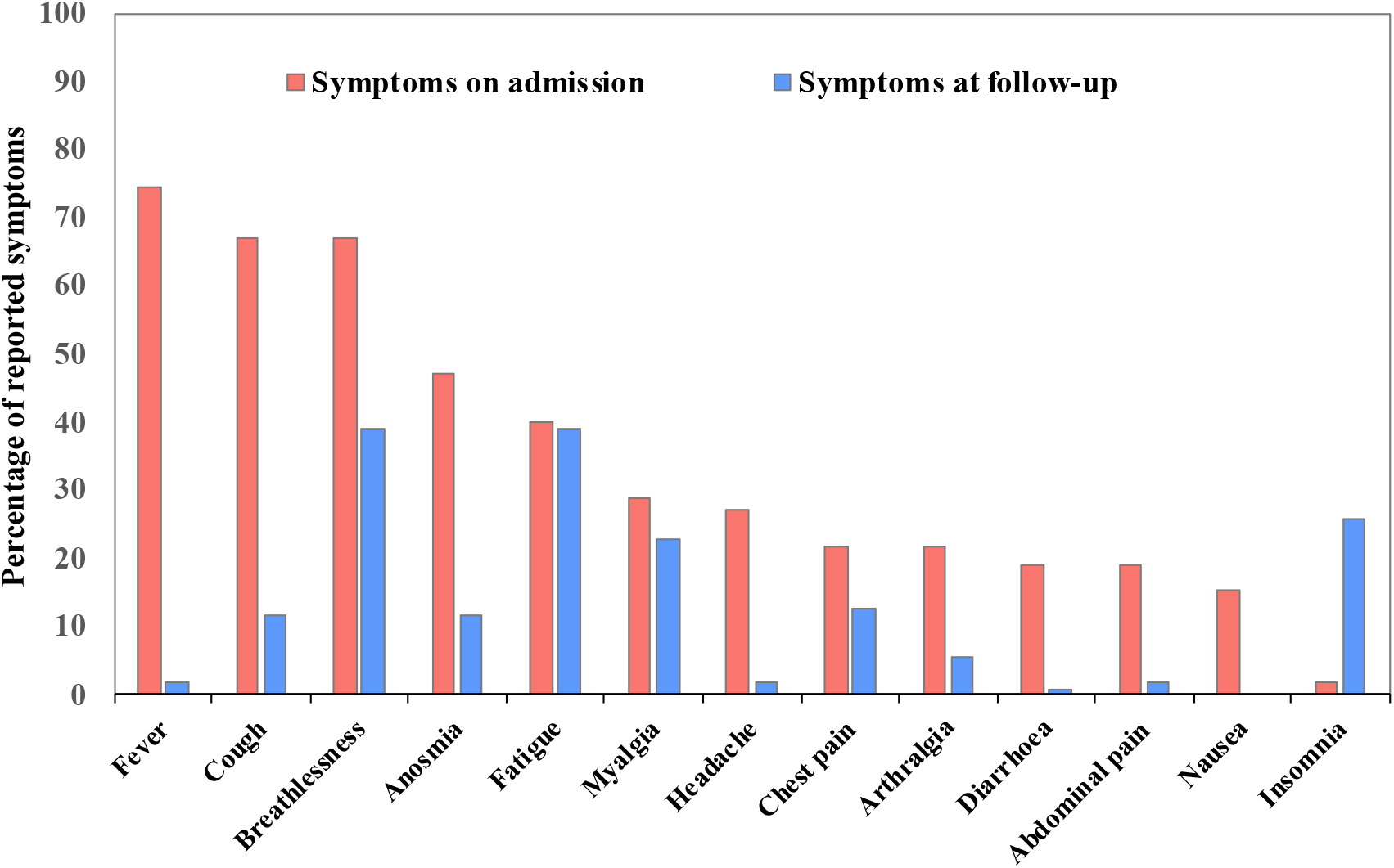
Frequency of symptoms reported at 12-week follow-up compared to hospital admission.

### Radiology

Of the 110 patients followed-up, 108 had a chest radiograph on admission (2 patients whom were pregnant did not have a baseline radiograph). The median baseline radiographic severity scores are described in Table 1. All participants had a chest radiograph performed at follow-up, with the vast majority (95/110) reported as normal. Of the 15 patients with abnormal radiographs (10 in Moderate group and 5 in Severe group), 2 had worsened from hospital admission with higher radiographic severity scores (both patients had known previous interstitial lung disease). Findings seen included consolidation (n=1) reticulation (n=8), atelectasis (n=5), pleural effusion (n=1). Nine high resolution computed tomography (HRCT) scans were performed on the basis of the clinical, spirometric or radiological findings, of which 2 showed evidence of fibrotic changes in patients with moderate disease at baseline. Both patients had restrictive pattern spirometry and had moderate COVID-19 disease during hospital admission.

### Pulmonary function testing

Baseline saturations were the same across all groups, as was baseline respiratory rate, see Table 2. Forced vital capacity as % predicted was lower in more severe disease. Eleven patients had restrictive pattern spirometry and 15 patients had a significant desaturation on STS test, all within the severe or moderate group.

**Table 2:** Spirometry and Sit-to-Stand desaturation test results

|  | Mild (n = 27) | Moderate (n = 65) | Severe (n = 18) | p-value |
| --- | --- | --- | --- | --- |
| O2 Saturations (%) | 98.0 (96.5, 99.0) | 97.00 (96.0, 98.00) | 97.0 (96.0, 98.0) | 0.88 |
| Nadir of O2 saturations on STS test | 96.0 (95.0, 97.0) | 95.0 (93.0, 96.5) | 95.0 (91.8, 96.0) | 0.75 |
| Respiratory rate | 17.0 (14.0, 18.0) | 17.0 (14.2, 19.8) | 17.0 (16.0, 18.0) | 0.95 |
| FVC (L) | 3.58 (3.13, 4.31) | 3.52 (2.75, 4.36) | 3.65 (2.55, 4.14) | 0.70 |
| FVC (% predicted) | 97 (90, 105) | 91 (78, 100) | 89 (76, 98) | 0.05* |
| FEV1 (L) | 2.97 (2.56, 3.42) | 2.71 (2.12, 3.49) | 2.54 (1.88, 3.23) | 0.50 |
| FEV1 (% predicted) | 94 (82, 101) | 90 (78, 100) | 89 (73, 101) | 0.30 |
| Restrictive pattern | 0 (0%) | 8 (12%) | 3 (17%) | 0.03* |
| Severe desaturation on STS test | 0 (0%) | 10 (15%) | 5 (28%) | 0.02* |
| *STS- 1 minute Sit to Stand desaturation test, FVC- Forced Vital Capacity, FEV1- Forced expiratory volume during first second of expiration.* |  |  |  |  |

### Health-related quality of life

SF-36 scores demonstrated a reduction in reported health status across all domains compared to population norms, see Figure 3. More severe COVID-19 disease at baseline was associated with greater deficiencies. In particular, physical scores were such as physical role and the composite physical score were significantly lower in the severe cohort, see Supplementary material for full results.

**Figure 3:**
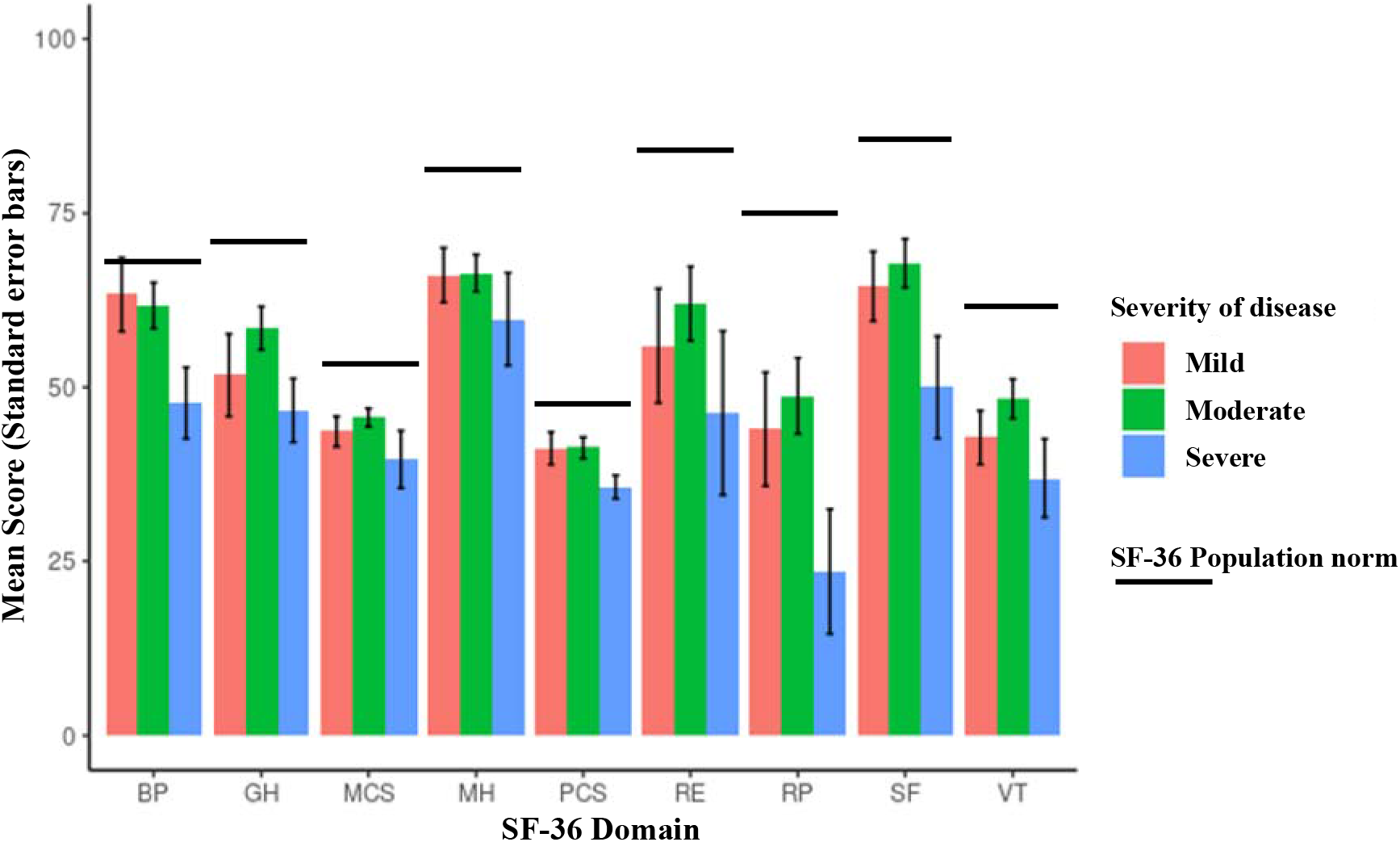
SF-36 results, mean and standard error, with age-matched population norm means.

There was no difference between the groups using the WEMWBS questionnaire (p=0.6 Kruskal-Wallis test). Medians for the mild, moderate and severe groups were 52 (IQR 44-56), 53 (IQR 42-59) and 50 (39-58) respectively. Results were similar to healthy population norms for this questionnaire.^18^

### Blood results

Thirty-five patients had significantly deranged liver or renal function recorded during their admission (9 renal, 19 liver). At follow-up, all had improved, and 32/35 results had returned to baseline. Across the cohort, 4 additional patients had significantly abnormal blood results including ongoing lymphopenia (n=2), CRP greater than 10mg/L (n=2). There was no difference between abnormal results and severity of disease, see summary of all results in Figure 4.

**Figure 4:**
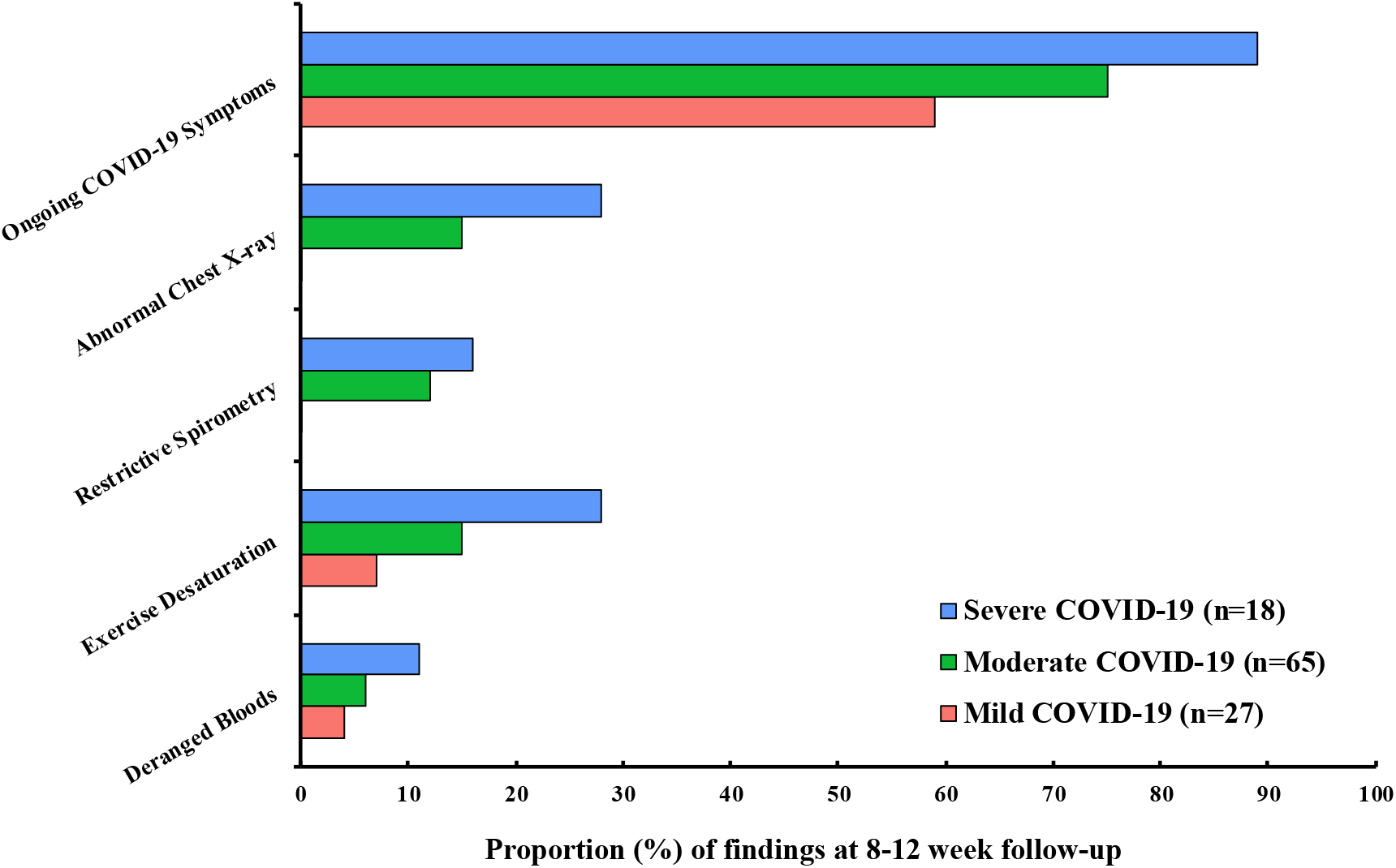
Summary of symptomatology and clinical results by disease severity.

## INTERPRETATION

Within the UK over 130,000 people have been admitted to hospital with COVID-19. As admission rates begin to fall the potential impact of “post-COVID” syndromes on patients and the health services is becoming apparent. The lack of prospective studies related to outcomes following COVID-19 has led to uncertainty as to how both hospitalised and non-hospitalised patients should be followed up and supported in their recovery. We present the first UK cohort study of consecutively recruited patients hospitalised with COVID-19 and systematically assessed after discharge.

One the most striking findings from this study is the persistence of symptoms relating to COVID-19 many weeks after the onset of symptoms or hospitalisation despite improvement in clinical and radiological parameters such as chest radiograph and blood results. Across the cohort, nearly three-quarters of patients had ongoing symptoms on questioning. Shortness of breath and excessive fatigue were the most predominant, but there were many others including chest pain, cough, fevers, arthralgia, myalgia, insomnia, dizziness. Even in patients who did not require oxygen in hospital and are therefore similar to many symptomatic individuals who may have self-cared or been managed by primary care, 59% had ongoing symptoms. In comparison, Marrie et al found 64% of patients hospitalised with pneumonia had symptoms at 6 weeks, and that time from pneumonia was a strong predictor of resolution.^19^ Given our patients were followed up a median of 12 weeks later and symptomatology was greater in COVID-19, this deserves further exploration.

Health related quality of life scores were lower in patients with more severe COVID-19 with particular deficits in patients perceived ability to perform their physical role and vitality. In contrast, there was no difference between groups reported mental wellbeing scores which were similar to UK population norms.^18^

The persistence of symptoms following COVID-19 infection is widely reported in the media and individual case reports but few studies have reported results from the routine follow up of cases post hospitalisation. In a study of patients discharged from an Italian hospital at a median of 60 days post discharge, patients were asked to recall their admission symptoms and HRQoL. It found that 87% had at least 1 ongoing symptom with fatigue (53%) and shortness of breath (43%) predominating, as in our study.^20^ Huang and colleagues assessed 57 patients approximately 30 days after discharge with pulmonary function testing. Despite the short timeframe post-discharge they detected abnormal spirometry in only a small proportion of the cohort (10% had low FVC and 9% had low FEV1).^21^ Due to different admission/discharge protocols in Chinese hospitals this cohort was much less severe at baseline than patients hospitalised in the UK. The cohort is much younger, required less oxygen with none documented as requiring invasive or non-invasive ventilation.

The lack of prospective follow-up studies on patients hospitalised with COVID-19 has resulted in guidance based on expert opinion and previous coronavirus infections. The British Thoracic Society (Guidance on Respiratory Follow Up of Patients with a Clinico-Radiological Diagnosis of COVID-19 Pneumonia published on 11th of May 2020) dichotomise follow up guidance depending on whether the patient required intensive/higher care versus ward/community care (equivalent to Severe versus Mild/Moderate in this cohort).^5^

For Mild/Moderate disease it recommends virtual follow-up with a chest radiograph and only proceeding to pulmonary function tests and further imaging if the chest radiograph is abnormal. This study demonstrated that 10% (10/92) of these patients would have an abnormal chest radiograph and all were considerably improved from hospital discharge. We would question the utility of routinely performing a chest radiograph in patients who did not require oxygen for their acute infection (unless an alternative pathology is suspected) as the likelihood of an abnormal result was zero in this cohort. We would support the approach of more intensive follow up in the severe group given a higher frequency of abnormal radiology and spirometry as well as a higher symptom burden.

A more intensive follow-up schedule has been recently published in Lancet Respiratory Medicine for all COVID-19 survivors regardless of severity of initial illness.^6^ In the first 12 months, 7 interactions with healthcare professionals (4 face-to-face) are recommended, alongside 4 HRCTs, 4 6MWT, 4 blood tests (including blood count and metabolic panel) and 2 SARS-CoV-2-IgG tests. Our prospective study of hospitalised patients does not support this approach outside of a research setting especially in patients with less severe COVID-19 at baseline, as the likelihood of detecting actionable spirometry and/or radiological findings is minimal. The prevalence of lung fibrosis post infection in this cohort was just 2% (2/110), far less than estimates following Severe Acute Respiratory Syndrome (SARS) or Middle East respiratory syndrome coronavirus (MERS-CoV).^8 9^ Extensive clinical investigations are not without harm, and follow-up research studies are planned which may be better placed to assess the role of intensive follow up.^22^

The lack of correlation between clinical test results and patients’ symptomatology and HRQoL has revealed an important aspect of follow-up which is advocated by the BTS guidelines; ‘Post-COVID holistic assessment’ in all groups including management of breathlessness and anxiety. All patients in these follow up clinics were offered referral to specialist psychological support services. Interestingly, the majority of patients were reassured by the knowledge of normal test results; just 5 patients were referred to specialist NHS psychology services.

To our knowledge this is the first study to prospectively recruit consecutive patients presenting with COVID-19 to hospital and systematically assess their outcomes after discharge. There remain several weaknesses which affect the generalisability of the study findings. Firstly, this is a single-centre study with relatively small patient numbers so rarer complications from COVID-19 may have been missed. Secondly, patients were followed up in a manner that might be replicated across may different hospital sites so cross-sectional imaging or full pulmonary function testing was not used routinely. At a time where waiting lists for such investigations are long and departments limited by personal protective equipment requirements, the availability of these tests are limited and should be used only when indicated.

In summary, this is the first follow-up study of consecutively consented hospitalised patients with COVID-19. The study demonstrates the persistence of symptoms at 12 weeks in the majority of patients, even those with milder disease initially. There was a reassuring improvement in clinical tests with only a minority having abnormal biochemical, radiological or spirometric tests. These results should inform the follow-up of post-hospital and community survivors of COVID-19 in primary and secondary care and shows that routinely performed clinical tests are unlikely to explain ongoing symptoms so holistic support of survivors is essential. Future studies should focus on the role of rehabilitation and psychological services for suffers of ongoing COVID-19 symptoms.

## Data Availability

Data available on reasonable request

## Authors and contributors

DTA, SB, NAM and FWH generated the research question and analysis plan. AM, AM2, MA, AN, CH, AB, EM, HA, JD, NAM, DTA, FWH were involved in data collection and clinical appointments. SG, JH, SH and KTE were involved in data analysis. All authors were involved in the final manuscript preparation.

## Funding

The DISCOVER study was supported by donations to Southmead Hospital Charity (Registered Charity Number: 1055900)

## Acknowledgement

The authors would like to thank the University of Bristol UNCOVER group for guidance in data analysis and study design.

## Notes

### Competing Interest Statement

The authors have declared no competing interest.

### Author Declarations

Ethics approval via South Yorkshire REC: 20/YH/0121

